# The decay of coronavirus in sewage pipes and the development of a predictive model for the estimation of SARS-CoV-2 infection cases based on wastewater surveillance

**DOI:** 10.1101/2022.03.16.22272521

**Authors:** Songzhe Fu, Qingyao Wang, Fenglan He, Can Zhou, Jin Zhang, Wen Xia

**Affiliations:** College of Marine Science and Environment, Dalian Ocean University, Dalian, China; Key Laboratory of Environment Controlled Aquaculture (KLECA), Ministry of Education; The Collaboration Unit for Field Epidemiology of State Key Laboratory for Infectious Disease Prevention and Control, Nanchang Center for Disease Control and Prevention, Nanchang, China

**Keywords:** Avian infectious bronchitis virus, SARS-CoV-2, Shedding route, Wastewater, Wastewater surveillance, decay, COVID-19

## Abstract

Wastewater surveillance serves as a promising approach to elucidate the silent transmission of SARS-CoV-2 in a given community by detecting the virus in wastewater treatment facilities. This study monitored the viral RNA abundance at one WWTP and three communities during the COVID-19 outbreak in the Yanta district of Xi’an city from December 2021 to January 2022. To further understand the decay of the coronavirus in sewage pipes, avian infectious bronchitis virus (IBV) was seeded in two recirculating water systems and operated for 90 days. Based on the viral abundance in the wastewater of Xi’an and the above data regarding the decay of coronavirus in sewage pipes, Monte Carol simulations were performed to estimate the infectious cases in Xi’an. The results suggested that the delta variant was first detected on Dec-10, five days earlier than the reported date of clinical samples. SARS-CoV-2 was detected on December 18 in the monitored community two days earlier than the first case and was consecutively detected in the following two sampling times. In pipelines without biofilms, the results showed that high temperature significantly reduced the viral RNA abundance by 2.18 log_10_ GC/L after experiencing 20 km travel distance, while only a 1.68 log_10_ GC/L reduction was observed in the pipeline with a low water temperature. After 90 days of operation, the biofilm matured in the pipeline in both systems. Reductions of viral RNA abundance of 2.14 and 4.79 log_10_ GC/L were observed in low- and high-temperature systems with mature biofilms, respectively. Based on the above results, we adjusted the input parameters for Monte Carol simulation and estimated 23.3, 50.1, 127.3 and 524.2 infected persons in December 14, 18, 22 and 26, respectively, which is largely consistent with the clinical reports. This work highlights the viability of wastewater surveillance for the early warning of COVID-19 at both the community and city levels, which represents a valuable complement to clinical approaches.

## Introduction

Severe acute respiratory syndrome coronavirus 2 (SARS-CoV-2) has caused a global pandemic and is still a public health emergency of international concern. Epidemiological surveillance of this virus has heavily relied on individual testing of clinical samples by quantitative real-time polymerase chain reaction (RT–qPCR). However, this approach was restricted by several factors, such as the inability to track asymptomatic disease carriers and reporting bias. Instead, wastewater surveillance has been proven to be feasible for the early warning of enteric viruses, such as norovirus, hepatitis A virus, and poliovirus, historically [1, 2]. Recently, wastewater surveillance has also been extensively used for the detection of SARS-CoV-2 in communities, as collected wastewater contains viruses excreted from both symptomatic and asymptomatic individuals in a certain catchment. Several recent studies that analyzed SARS-CoV-2 titers in wastewater treatment plants (WWTPs) by RT-qPCR revealed a good correlation between SARS-CoV-2 incidence rates and virus titers in wastewater [3-7]. Ahmed et al. (2020) further established a model to estimate the number of infected individuals based on the concentration of viral RNA; the model estimated a median range of 171 to 1,090 infected persons in the catchment, which is in reasonable agreement with clinical observations [7]. Recent studies also showed the additional value of wastewater surveillance for the detection of SARS-CoV-2 before they are reported by local clinical tests[8, 9]. There is a need to understand the persistence of SARS-CoV-2 and its nucleic acid in water and wastewater environments to properly interpret SARS-CoV-2 RNA measurements from wastewater collection systems.

The sensitivity of the analytical method is very important in tracking SARS-CoV-2 in wastewater. However, the transportation of wastewater in municipal pipelines with biofilms and the use of chemicals such as chorine dioxide would largely affect the variability of viruses. In addition, high water temperature would also result in the decay of the virus. The detection might be negative for viruses at low concentrations, which would underestimate the silent transmission of SARS-CoV-2 in the community. To date, no study has elucidated the mechanisms influencing the transport of viral RNA in sewage pipelines. Studies are urgently needed to assess viral decay due to retardation of its transport due to sorption on biofilm and the pipe network material as well as chemical inactivation. The outcome of these studies will enable better prediction of the viral RNA level at the source. Continuous sampling along the sewerage pipeline with known flow rates would aid in the evaluation of the decay of the virus and facilitate more accurate tracking of the virus concentration.

In this study, we first evaluated the decay of a gamma coronavirus (CoV) during the transportation of wastewater, which was used for the development of a predictive model for the estimation of SARS-CoV-2 infected persons. Next, we conducted continuous wastewater surveillance in Xi’an, a northwest city in China. Our initial purpose was to monitor the presence of Hanta virus (causing epidemic hemorrhagic fever) by wastewater surveillance. Unexpectedly, SARS-CoV-2 was monitored in three communities and one WWTP that recently witnessed a large COVID-19 outbreak.

## Materials and Methods

### Effect of transportation of wastewater on viral activity

In this study, to analyze the virus decay along the long-distance municipal pipeline, a recirculating water system was set up. The system consists of two 80 m^3^ tanks (with 60 m^3^ of synthetic wastewater) and a pipeline loop with a total length of 100 m. The synthetic wastewater was prepared as described previously [10]. A mixture of gelatin, polyoxyethylene-sorbitan monooleate, and starch was prepared to simulate the polysaccharide, protein, and lipid components of municipal wastewater. The time required for one circulation is approximately 15 mins. Briefly, wastewater was seeded with 10^6^ GC/L IBV (final concentration). Once the IBV was seeded into the tank, it would travel 9600 m in 24 h. We sampled 200 ml wastewater when the travel distance reached 5, 10, 15, and 20 km (sampling at 12.5, 25, 37.5, and 50 h, respectively) to evaluate the effect of transportation of wastewater on the viral activity. Once the experiment was finished, the inoculated wastewater was disinfected by dioxide chlorine for 60 mins before discharging into the sea. Meanwhile, to evaluate the effects of biofilms on the viral activity in the municipal pipeline, IBV was also seeded into the two systems on day 60. Wastewater was also sampled at the above travel distance and discharged into the sea after 50h.

### Biofilm bacterial count analysis

To count the sessile bacteria in the biofilm, 5 cm × 5 cm areas in the internal pipeline were swabbed and gently washed with 0.1 mol/L sterile PBS to remove planktonic bacteria. Afterward, the samples were placed into a 5 mL sterile PBS tube and then vibrated with a vortex meter for 60 s to remove the biofilm. Subsequently, 0.1 mL biofilm bacterial solution was diluted with PBS from 10^−2^ to 10^−6^. Finally, the number of sessile bacteria was determined by spreading onto 2216E marine agar.

### Calculations of decay rate of IBV in wastewater pipeline

IBV RNA concentrations (GC/L) at various temperatures or different travel distances were used to calculate the decay rate. The observed IBV RNA concentrations were linearized using the natural log (ln)-transformation of the IBV RNA concentrations, as shown in Equation (1) [11]. These values and their associated travel distances were used to calculate the first-order decay rate constants in units per kilometer by linear regression in the R package. The fit of the regression and the appropriateness of the linear model were assessed by the root mean square error (RMSE) and R^2^ value, which assesses the fit by measuring the distance of the observed values from the fitted line.

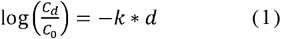

where C_0_ and C_t_ are the IBV RNA concentrations of GC in the wastewater at travel distances of 0 and d km, respectively, and k is the decay rate constant. The distance required to achieve a 90% (one log) reduction (D90) was calculated using equation (2).

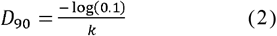

After the mean *k* values were estimated for each treatment, the values were log10-transformed, and linear regression was used to characterize the effects of different treatments on the first-order decay rate constant. Two-way ANOVA with Tukey’s multiple comparison tests and paired t tests were performed to evaluate the effect of treatment factors (biofilm matrix and temperature) on the decay of IBV RNA. All statistical analyses were performed in the R package, and a P value less than 0.05 was considered statistically significant.

### Clinical data collection

Data on COVID-19 infection in Xi’an were obtained from the National Health Commission of the People’s Republic of China released in public between 9 December, 2021, and 20 January, 2022. The information includes the demographic, epidemiological, and travel trajectory of individual cases. A previous study suggested that the majority of viral shedding from the feces occurred 1-5 days after the infection, regardless of COVID-19 severity [12, 13]. Thus, we estimated the actual number of infection cases as the confirmed number of cases plus the cases detected in the next

1-5 days.

### Sewage samples and pasteurization

We collected wastewater on 10 December, 14 December, 18 December, 22 December, and 26 December of 2021 and on 15 January 2022 from the influent of a WWTP in Yanta district of Xi’an city and a manhole in three communities. To reduce the variability of grab sampling, 200 ml of sewage samples were collected in the morning peak (8 am to 10 am) at 15-min intervals for 2 h and pooled together. Upon initial receipt, samples were placed in the biosafety cabinet with UV for 20 min and then pasteurized in a 60°C water bath for 60 min to inactivate the virus. Pasteurized samples were then used for viral precipitation, and the remaining samples were stored at 4°C.

### RNA extraction

RNA extraction was performed as described by Wu et al. (2020)[14]. Briefly, the wastewater samples (100 mL) were first centrifuged at *4750 g* for 5 mins. The supernatant was mixed with 8.0 g of PEG8000 and 4.7 g of sodium chloride to final concentrations of 10% and 1 M, respectively. The mixture was incubated at 4°C overnight with 100 rpm agitation, followed by centrifugation at 10,000 g for 2 h at 4°C to pellet the virus particles. The pellet was then resuspended in 200 µl of phosphate buffered saline (PBS). RNA was extracted using QIAamp Viral RNA Mini Kits (Qiagen) according to the manufacturer’s instructions with a final elution volume of 50 μL. Meanwhile, IBV obtained from chicken feces from a previous study [15] was used as a molecular process control to monitor the efficiencies of the RNA extraction and RT–qPCR processes. Briefly, 150 μL of the virus concentrate was spiked with 2.1 × 10^5^ copies (2 μL of a viral stock) of IBV and subjected to RNA extraction as described above.

SARS-CoV-2 ORF1ab gene and Hanta virus were detected by two commercial RT–qPCR detection kits, respectively (Shanghai Zhijiang, Shanghai, China). IBV was detected by another commercial detection kit (Shanghai Xinyu Biotech, China). RT–qPCR amplifications were performed in 40 μL reaction mixtures using iTaq™ Universal Probes One-Step Reaction Mix (Bio–Rad Laboratories, Richmond, CA). All RT-qPCRs were performed in triplicate. For each qPCR run, a series of three positive and negative controls were included.

### Monte Carol simulation to estimate the infection number in the community and district

Although individual shedding in saliva, sputum, stool, and urine all contribute to the virus load in wastewater, herein we only examined the probable contributions of stool and urine to the virus load of wastewater, as a previous study suggested that the virus loads in saliva and sputum are 3 log_10_ lower than those in stool and urine[16].

The amount of SARS-CoV-2 RNA copies/g in feces was modeled as a log-uniform distribution from 2.56 to 7.67 log_10_ copies/g, as observed in a previous study [17]. The daily stool mass per person was modeled as a normal distribution with a mean of 211 g as reported by Rose et al. (2015)[18]. Meanwhile, we assumed that the virus load in urine followed a distribution with a mean of 2.91 log_10_ copies/ml in urine and an average urination amount of 1500 ml (Table S1). Based on the data from Xi’an in 2020, the average water use is 135 L/person/d.

The sampled WWTP in Yanta district has a catchment of approximately 900,000 persons (capita). The observed average per capita wastewater rate in this region was 135 L/person/day in 2021. We first used Eq. (1) to obtained the corrected SARS-CoV-2 RNA copies without decay:

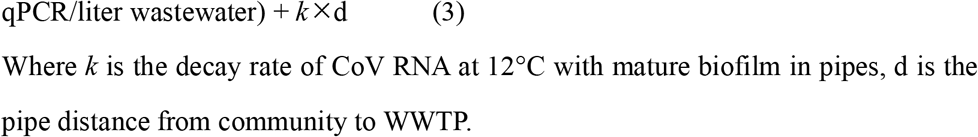

Where *k* is the decay rate of CoV RNA at 12°C with mature biofilm in pipes, d is the pipe distance from community to WWTP.

Then, we used the above assumed virus load in stool and urine as input parameters and considered the virus decay in the pipes, and the predicted number of persons infected can be calculated by the following equations (4) (WWTP model):

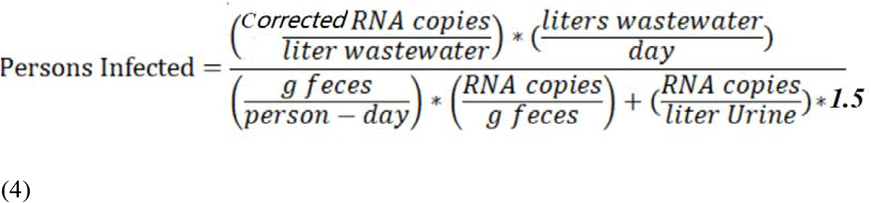

For a single community, the virus load will be first diluted in the community. The dilution factor equals the number of individuals in the communityX daily water use per person. Thus, the infection cases can be modeled by the following equation (5) (community model):

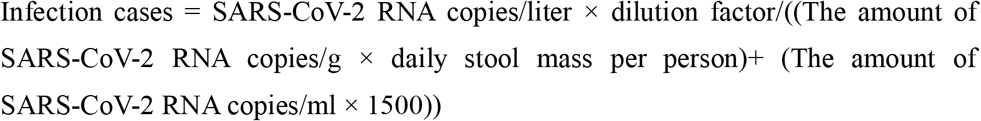

The Monte Carol simulation was built in Excel (Microsoft, Redmond, WA) with ModelRisk version 6.0. The number of iterations per simulation was 10,000. The root mean square error (RMSE) was used as a criterion for selecting probability distributions based on goodness of fit. Maximum population densities estimated in the simulations. Sensitivity analyses were graphically represented by tornado charts [19]. Plots were automatically generated in ModelRisk by selecting predicted infection cases as the output. For the tornado plot, a rank order correlation was used, which provided a statistical measure of the correlation between the five most important inputs and output generated values. The predicted infection cases are reported as the median and 95% confidence interval (CI) determined by bootstrapping the model with 100 experiments of 1,000 draws each.

## Results

### Effect of transportation of wastewater on viral activity

To evaluate the effect of transportation of wastewater on viral activity, we seeded IBV in the influent of two recirculating systems. The water temperature of the two systems was monitored throughout the study, with the temperatures maintained at 12.0 ± 2°C and 28.0 ± 2°C throughout the duration of the experiment. The mean concentrations of IBV RNA in the two studied pipelines at day 0 ranged from 6.03 ± 0.20 and 5.84 ± 0.13 log10 GC/L, respectively. The declining concentrations of IBV RNA at travel distances of 5, 10, 15, and 20 km at 12 and 28°C are shown in Fig. 2. The log10-transformed mean decay rates for IBV RNA with or without biofilm at two temperatures observed in the study are shown in Table 1. For the no biofilm pipeline, the mean first-order decay rate constants (*k*) were 0.0799/km (R^2^ =0.957) at 12°C to 0.12/km at 28°C (R^2^ =0.957) (Table 1). Within the 5 km travel distance, there was no statistically significant decay of IBV in low-temperature wastewater, while over a 1.68 log decline of IBV was observed when the travel distance reached 20 km. In high-temperature wastewater, significant decay was observed at a 5 km travel distance, and an over 2.18 log reduction in IBV was found at a 20 km travel distance. The average IBV RNA D90 (distance required for one log_10_- reduction) was 12.51 and 8.32 km for experiments conducted at 12°C and 28°C, respectively.

**Table 1.**
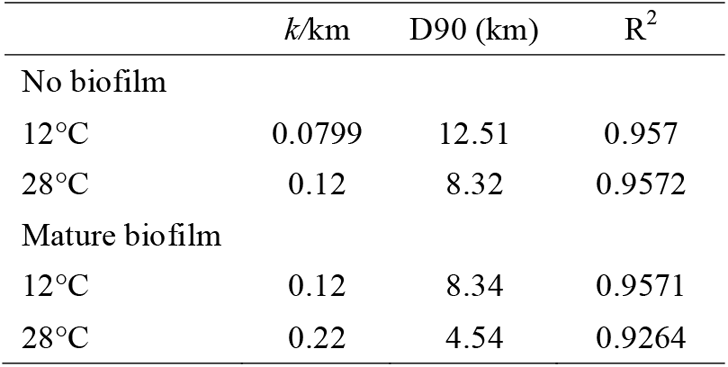
Decay rate (*k*/km) and *D*90 values of avian infectious bronchitis virus (IBV) RNA in wastewater pipelines with or without biofilms

**Figure 1.**
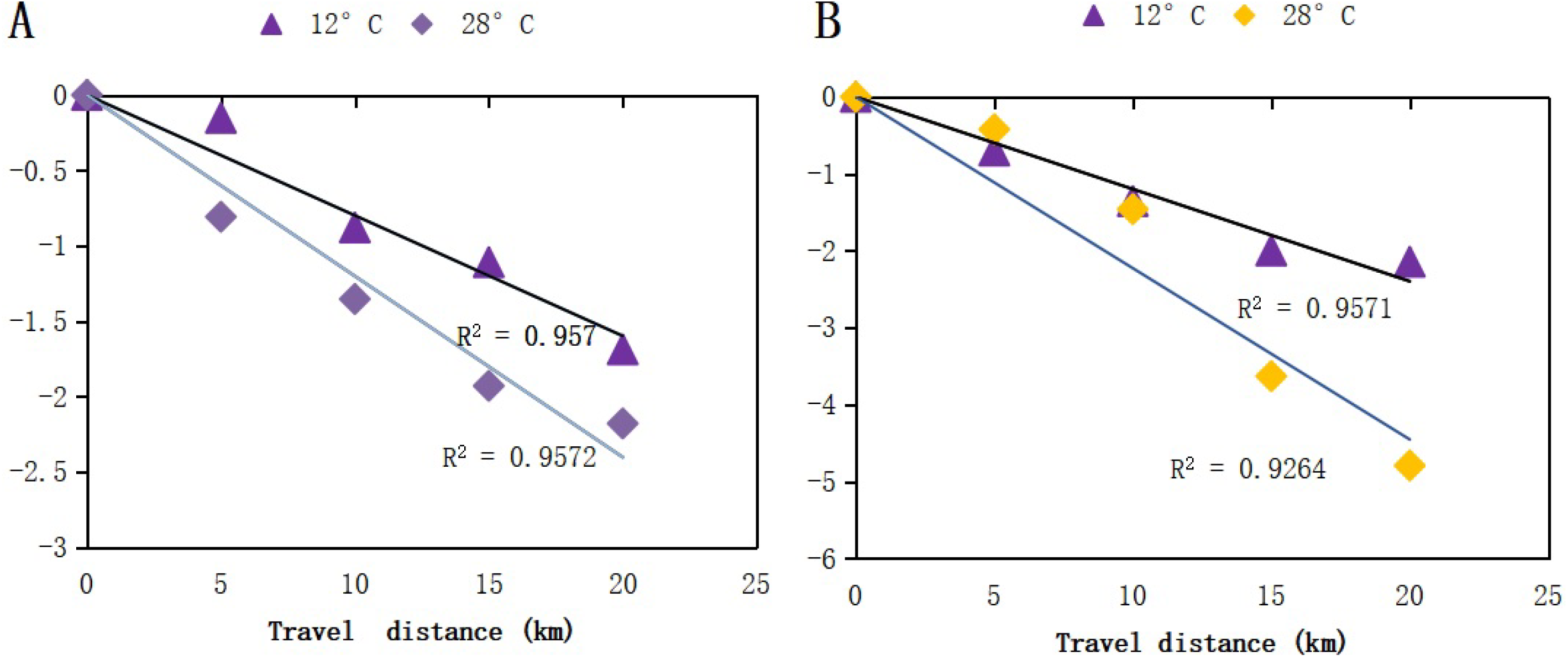
Mean decay curves of IBV RNA over travel distance (km) in untreated wastewater pipeline without (A) or with biofilm (B). The measurements were linearized premised on first-order decay, in which the natural log (ln)-transformed measured concentration at each time point was divided by the concentration at time zero. The error bars (SD) are too small to illustrate.

**Figure 2.**
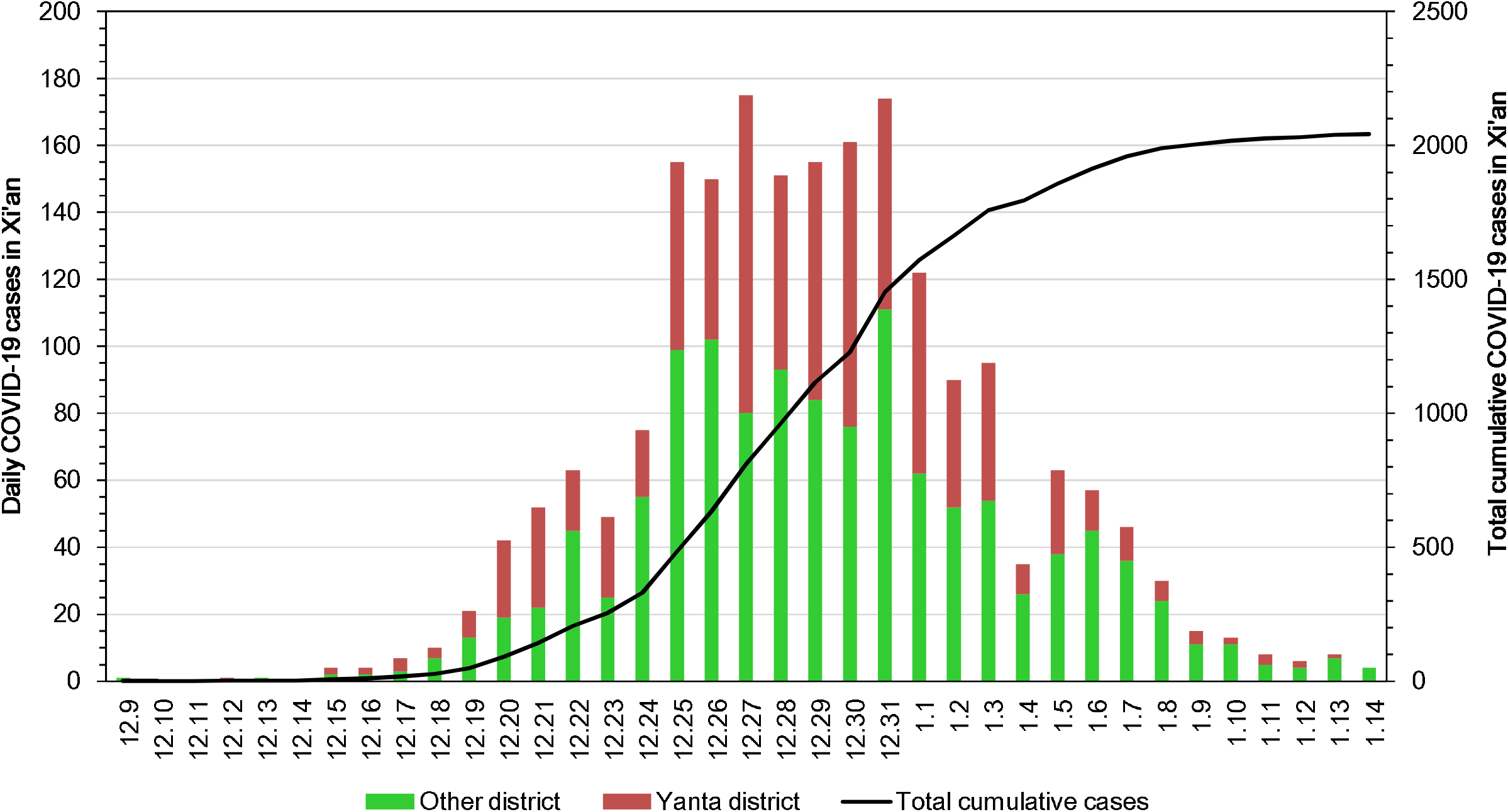
Daily COVID-19 cases in Xi’an city and Yanta district from December 9, 2021, to January 15, 2022.

### Effect of biofilm matrix in the pipeline on viral activity

Next, we further examined the effect of the biofilm matrix in the pipeline on IBV decay when it passed through the pipeline. We first evaluated biofilm formation during the experiment. For low-temperature wastewater, the number of sessile bacteria on day 0 for the two systems averaged 11 cfu/cm^2^. With increasing time, the bacterial counts in the biofilm in the high-temperature system increased from 4.79 to 7.92 log_10_ cfu/cm^2^ on days 15 and 45, respectively, and then remained stable until day 90 at approximately 7.9 log_10_ cfu/cm^2^. This suggests that a mature biofilm was formed after 45 days. For low-temperature wastewater, the number of sessile bacteria increased from 3.53 log_10_ cfu/cm^2^ on day 15 to 5.19 log_10_ cfu/cm^2^ and reached a plateau on day 60. The above observations suggested that for both low- and high-temperature wastewater, biofilms would be mature after 60 days. As most municipal pipelines have more than 60 days of operation, we sampled wastewater at day 60 to examine the effect of the biofilm matrix in the pipeline on the viral activity for both systems. In high-temperature wastewater, decay rate constant was 0.22/km (R^2^ =0.95) and was greater than the constant for wastewater at low temperatures (*k*=0.12/km). The estimated IBV RNA D90 was 8.34 km at 12°C and 4.54 km at 28°C.

### Epidemiological description of the Xi’an COVID-19 outbreak

On the 9^th^ December, one person was confirmed to be SARS-CoV-2 positive in Xi’an city. Subsequent mass testing registered a massive outbreak of COVID-19 in the following 40 days, with a total of 2053 cases in this city. Infection cases were found in 13 districts of Xi’an city, of which 40.26% were found in Yanta district.

The infection case in Yanta district was initially found in Dec 12, and daily infection cases gradually increased from 2 cases in Dec 15 to 95 cases in Dec 27. Afterward, the number of reported cases gradually decreased and reached zero on Jan 14. For the sampled community, a total of five cases were identified in Community One, which were diagnosed on Dec 20, 22, 25, 29 and 31, respectively. No SARS-CoV-2 was detected in Community Two and Three.

### Detection of SARS-CoV-2 RNA in wastewater samples

From Dec 1^st^, 2021, in the Yanta district, we monitored SARS-CoV-2 in the inlet municipal wastewater of a WWTP and a manhole in three communities in this district. Wastewater surveillance was conducted on Dec 1^st^, Dec 5^th^, Dec 14^th^, Dec 18^th^, Dec 22^nd^, Dec 26^th^, Dec 31^st^ and Jan 15^th^. The first SARS-CoV-2-positive water sample was detected on Dec 14^th^ with an average virus load of 3.83×10^2^ GC/L for ORFab1 genes; at that time, two clinical COVID-19 cases attributed to the delta variant had been identified in Yanta district, but none from the communities. Samples collected on Dec 18^th^, Dec 22^nd^, and Dec 26^th^ showed an ongoing outbreak of COVID-19 during this period, harboring average virus loads of 6.89×10^2^, 1.54×10^3^ and 4.56×10^3^ GC/L, respectively. In the monitored community, SARS-CoV-2 was first detected on Dec 18^th^ with an average virus load of 1.42×10^5^ GC/L; two days after the wastewater data were reported, one infection case had been identified in a resident of the community.

Samples collected on Dec 22nd and Dec 26th from this same sewershed also retrieved SARS-CoV-2 with concentrations of 1.42×10^4^ and 1.66×10^4^ GC/L, respectively.

Another two new cases were diagnosed on Dec 22 and 25. On Dec 26th, the close contactors of infection cases were transferred into the quarantine place, and two new cases were identified in the quarantine place. Afterwards, the concentration of SARS-CoV-2 dropped to a level below the detection limit until the end of the surveillance.

### Community model to estimate the actual infection cases

As summarized in Table 2, the Monte Carlo simulation estimates a median number of 3.68 (0.86-9.97, 95% CI) infected persons on Dec 18^th^ and 5 (1.13-13.68, 95% CI) on 22/12/2021 in the monitored community. This is consistent with the clinical observation; three cases were reported until Dec 22^nd^, while two additional cases were found four days later. The sensitivity analysis indicated that the estimated number of infections and prevalence were strongly correlated with the log_10_ SARS-CoV-2 RNA copies in stool, followed by the RNA copies detected in urine and the amount of feces/person/day. The model was least sensitive to the travel distance of wastewater (Figure S2).

**Table 2.** Number of SARS-CoV-2-infected persons and prevalence in the treatment catchment basin as estimated by viral RNA copy detection in wastewater and Monte Carlo simulation

| Sampling point | RNA copies/L | Estimated infection cases* | Predicted infection cases |
| --- | --- | --- | --- |
| WWTP |  |  | median (95% CI) |
| 14-Dec | 383.2 | 2-19 | 23.3 (5.33-63.83) |
| 18-Dec | 689.5 | 19-106 | 50.11 (11.4-139.62) |
| 22-Dec | 1542 | 114-238 | 127.3 (29.1-347.5) |
| 26-Dec | 4160 | 243-420 | 524.2(118.5-1436.1) |
| community one |  |  |  |
| 18-Dec | 14200 | 2 | 3.68(0.86-9.97) |
| 22-Dec | 16600 | 5 | 5 (1.13-13.68) |
| community two |  |  |  |
| 18-Dec | ND | 0 | NA |
| 22-Dec | ND | 0 | NA |
| 26-Dec | ND | 0 | NA |
| community three |  |  |  |
| 18-Dec | ND | 0 | NA |
| 22-Dec | ND | 0 | NA |
| 26-Dec | ND | 0 | NA |

\*including the detected cases in the next 1-5 days

### Prediction of the actual infection cases by the WWTP model

Meanwhile, based on previous studies, actual infection cases were calculated based on the reported cases on the wastewater sampling date plus the ones detected in the next 1-5 days. Thus, we estimated the actual infection cases on Dec 14^th^ (2-19 cases), Dec 18^th^ (19-106 cases), Dec 22^nd^ (114-238 cases), and Dec 26^th^ (243-420 cases). Given the uncertainty and variations of the input parameters, the WWTP model estimated a median value of 23.3, 50.11, 127.3, and 524.2 infected persons in the catchment on Dec 14^th,^ Dec 18^th,^, Dec 22^nd^ and Dec 26^th,^, respectively (Figure S3), which is in reasonable agreement with clinical infection cases at the above four time points.

### Detection of Hanta virus in wastewater samples

From Dec 1^st^, 2021, to Jan 15^th^, 2022, no Hanta virus was detected in wastewater samples at both WWTP and community levels.

## Application of predictive model in other cities

### 1. Detection and prediction of SARS-CoV-2 infection cases in Yingkou city

From April 2021 to April 2022 in the Bayuquan district, Yingkou, we monitored the SARS-CoV-2 in the untreated urban wastewater in Yingkou. Wastewater surveillance was conducted weekly for SARS-CoV-2 in the Bayuquan district beginning on the 20^th^ April (Table S4). The first SARS-CoV-2-positive water sample was detected on the 27^th^ April with CT values of 41.37 for ORFab1 genes to an average virus load of 0.43×10^1^ Genome Unit/100 ml. Samples collected from 5^th^ May, 12^th^ May and 20^th^ May evidenced an outbreak of COVID-19 during this period, harboring a virus load of 6.1×10^2^, 1.78×10^3^ and 2.29×10^1^ Genome Unit/100 ml, respectively. The concentration of SARS-CoV-2 dropped to a level below the detection limit on 5^th^ June. No SARS-CoV-2 was detected afterward until the end of surveillance.

On the 14^th^ May, one person was confirmed SARS-CoV-2 positive in the Bayuquan district. Subsequent mass testing registered an outbreak of COVID-19 in the following six days, with 14 new cases in the district (Figure 5A).

### 2. Detection and prediction of SARS-CoV-2 infection cases in Dalian city

From Dec 2020 to Nov 2021, we conducted annual wastewater surveillance on the influent of three WWTPs and one wastewater effluent in seashore in Dalian city, including Malan WWTP, Chunliu WWTP, Dagushan WWTP and Heishijiao

### 3. Detection and prediction of SARS-CoV-2 infection cases in Nanchang city Discussion

In this study, we investigated whether the presence of SARS-CoV-2 in untreated wastewater can be used as an early warning for COVID-19 infections in communities. The detection of clinical samples remains the gold standard for disease surveillance and tracking. However, such data are limited due to factors such as reporting bias and inability to track asymptomatic disease carriers. The advantage of wastewater surveillance is that infectious agents are excreted in the urine and feces of infected individuals regardless of disease symptom severity. Thus, it is a promising approach for early warning.

This study first monitored the presence of SARS-CoV-2 in three communities and one WWTP during a large-scale outbreak in Xi’an city. Expectably, SARS-CoV-2 was detected 4 days earlier than the clinical data were reported, suggesting that SARS-CoV-2 has circulated in the local communities before massive testing starting on Dec 18^th^.

However, at present, no study has investigated the survival of CoVs during their passage through the sewer pipe network, which affects the accurate estimation of infection cases via wastewater surveillance. Current evidence suggests that CoVs do not survive well in aqueous environments relative to noroviruses, which can persist for months[18]. Herein, we used IBV as a surrogate CoV to analyze its decay under different treatments. Our study examined the survival of IBV in a stimulated sewer pipe under different treatments, which provided critical information to understand the survival of CoVs during their passage through the sewer pipe network. Equations describing the linear regression fit for each treatment were summarized. The fit of each linear model was reasonable, with R^2^ values ranging from 0.927 to 0.957. As expected, the log10-transformed first-order decay rate increased with increasing temperature IBV RNA with or without the biofilm matrix. However, the decay rate increased more rapidly with increasing temperature in the pipeline having a mature biofilm, indicating that the first-order decay rate was also sensitive to increasing travel distance. This is consistent with a previous study. Ahmed et al. (2020) examined the decay rate of murine hepatitis virus and gamma-irradiated SARS-CoV-2[7]. Their results suggested that there was no statistically significant decay at lower temperatures (4°C and 15°C) irrespective of the virus type and water source, but the decay at higher temperatures (25°C and 37°C) was significantly faster than that at lower temperatures. Notably, their results also showed that SARS-CoV-2 RNA decayed faster in untreated wastewater than in tap water, indicating the role of the matrix in accelerating virus decay.

Another factor contributing to the degradation of SARS-CoV-2 is wastewater treatment, including filtration, sedimentation and disinfection. The results herein suggest that the levels of SARS-CoV-2 are greatly reduced by 3.1 log_10_ gc/ml during the disinfection of stimulated wastewater treatment (data not shown), suggesting that the virus is degraded during disinfection. This is consistent with studies showing a 2 to 3 log_10_ removal efficiency in viral RNA abundance when comparing viral levels in influent and effluent [20]. Combined with the above factors, we developed a modified Monte Carlo simulation for both communities and WWTPs to estimate the number of infection cases based on the viral RNA concentration in wastewater. Thus, we proposed a new road-map for the prevention of SARS-CoV-2 outbreak (Figure 3). Firstly, routine wastewater surveillance is conducted twice a week in WWTP to monitor the early signals of SARS-CoV-2. Once WWTP sample is positive, a emergent reaction will be triggered to sample the wastewater from community which should be finished in 24h. Meanwhile, the community would be blocked during this period. Afterwards, massive clinical test would be conducted in the communities with positive wastewater sample, while the shutdown of other communities can be canceled if both wastewater and antigen test are negative twice, which can minimize the disturbance of daily live.

**Figure 3.**
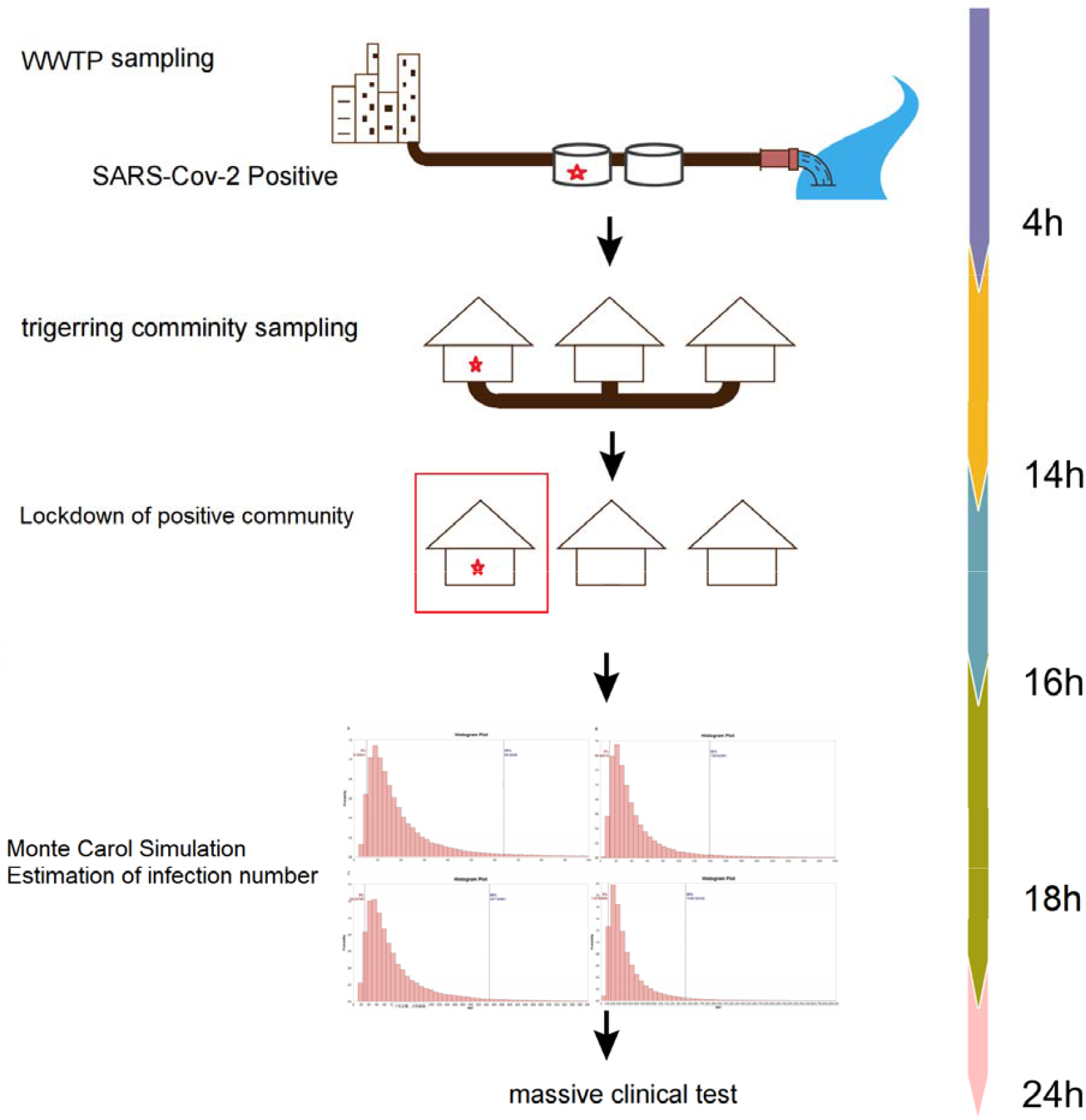
A new strategy for rapid estimation of SARS-CoV-2 infection cases based on wastewater surveillance

There are two limitations of this study. First, SARS-CoV-2 was replaced by IBV to understand the dynamics of CoV in the wastewater pipeline. Ideally, it would be much better to use intact SARS-CoV-2 seeding in the pipeline; however, this is not possible due to high biological safety level requirements for working with SARS-CoV-2 in a pilot-scale wastewater treatment plant. Therefore, a comparative study using SARS-CoV-2 as seeding material is needed to assure the reproducibility of the modeling. However, the results from Ahmed et al. (2020) showed that the decay rates of MHA and SARS-CoV-2 were comparable across all treatments with no significant difference, indicating the feasibility of using other CoVs for such analysis[22]. Nevertheless, comprehensive comparative studies are needed to reach a consensus on the surrogate virus used in modeling.

Another limitation is that there is also significant uncertainty and variations in the model input parameters, especially for the distribution of SARS-CoV-2 RNA concentrations in stool and urine and the volume of excreta produced to the sewer system. This paper is largely limited to currently available data, and future modeling with updated input parameters with more specific and representative data would further improve the accuracy. However, uncertainty and variations do not preclude the usefulness of the Monte Carol model to infer the person infected. Despite the high variability within and between individuals, both the community model and WWTP model achieved reasonable predictions for the infection cases.

## Conclusion

In conclusion, we conducted wastewater surveillance on a WWTP and three communities during the COVID-19 outbreak in Xi’an city. To better predict the infection cases, we examined the effects of wastewater transportation in the pipeline on the decay of CoV. The results suggested that the biofilm matrix and temperature appear to be important factors affecting the decay of IBV, resulting in the degradation of CoVs by up to a 4.8 log_10_ reduction. Next, a revised model was developed to predict the number of persons infected in the Xi’an outbreak. Our results clearly showed that most predictions are in reasonable agreement with clinical reports, highlighting a promising tool for the early warning of infectious diseases.

## Data Availability

All data produced in the present work are contained in the manuscript

## Acknowledgement

We thank volunteers in Xi’an for sampling assistance. National Natural Science Foundation of China (81903372) and Science and Technology Department of Jiangxi Province, China (20202BBGL73053) supported this research.

## Notes

### Competing Interest Statement

The authors have declared no competing interest.

### Summary of Updates

corrected some typo errors

