## Supplemental Figure 1-3 for "The decay of coronavirus in sewage pipes and the development of a predictive model for the estimation of SARS-CoV-2 infection cases based on wastewater surveillance"

Table S1 Fitted model parameters and sources for Monte Carol simulation

| Parameter | Unit | Distribution | Parameters | Number of Observations | Citation |
| --- | --- | --- | --- | --- | --- |
| transportation distance | kilometer | Pert | mean = 2.5, min =0.3, max = 12.3 | 203 | NA |
| GC SARS-Cov-2/gram stool | log_10_ GC/g | Pert | mean = 5.4, min = 1.9, max = 10.3 | 166 | (Han et al., 2020  ; Lescure et al., 2020; Wölfel et al., 2020) |
| stool production per day | g/day | Normal | mean = 211, min=0, max=520 | 357 | (Rose et al., 2015) |
| GC SARS-Cov-2/mL urine | log_10_ GC/mL | Triangle | mean = 3.31, min = 0.59, max = 7.55 | 15 | (Kashi et al., 2020; Roshandel et al., 2020; Yoon et al., 2020) |

Figure S1 Dynamics of bacterial counts in the biofilm of the wastewater pipelines at 12°C and 28°C from day one to day 90.

Sessile bacteria counts(log10 cfu/cm2)

| Immersion time (days) | 12°C | 28°C |
| --- | --- | --- |
| 0 | 1.01072 | 1.13437 |
| 15 | 3.53514 | 4.78813 |
| 30 | 4.75593 | 5.16668 |
| 45 | 5.19033 | 7.92601 |
| 60 | 5.8451 | 7.94565 |
| 90 | 5.81291 | 7.97978 |

**Figure S2** The sensitivity analysis based on tornado plot

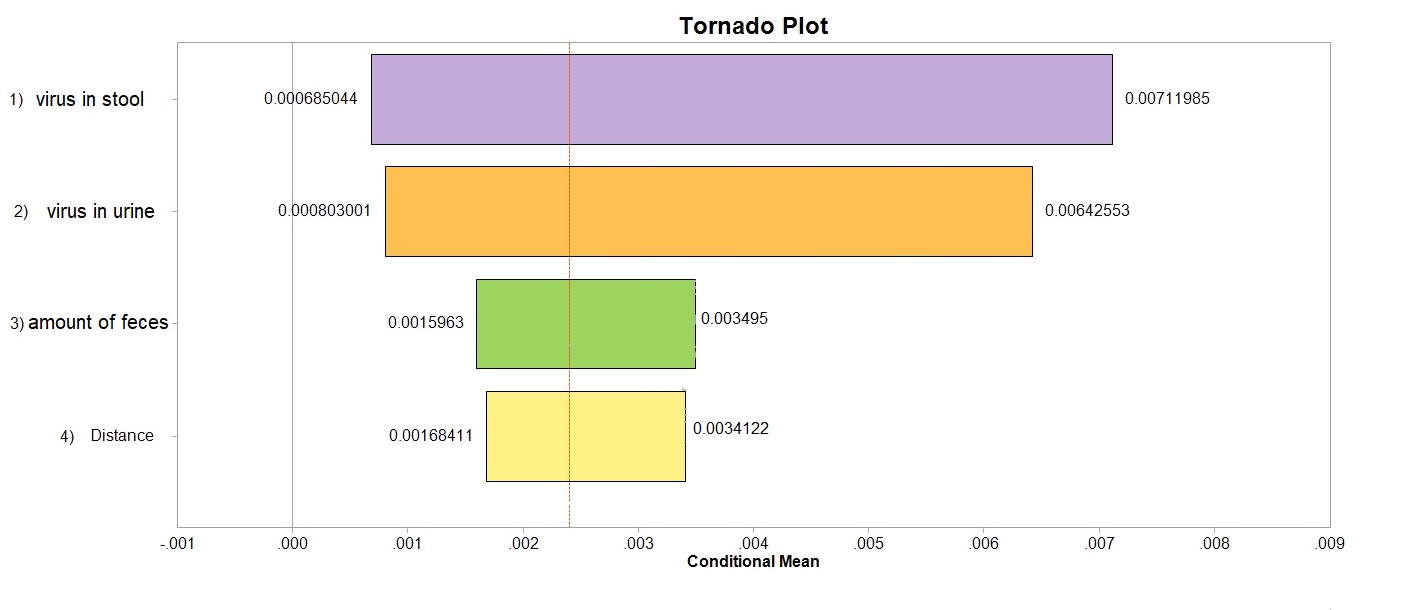

Figure S3 Histogram Plot of Monte Carol simulation results on Dec-14 (A), Dec-18 (B), Dec-22 (C), and Dec-26 (D).

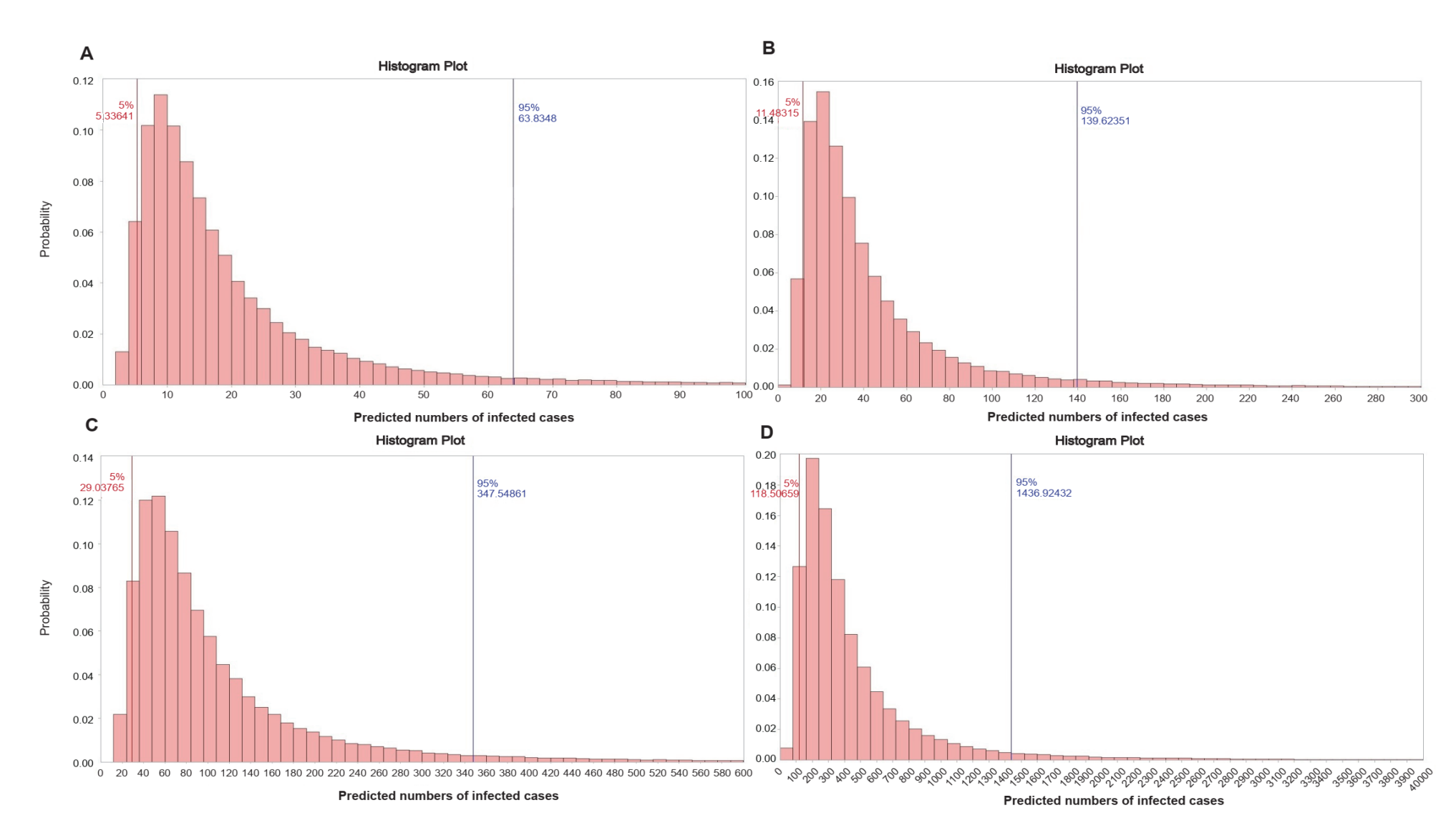
